# Social responsibility is the crucial factor in adopting early vaccination plans

**DOI:** 10.1101/2020.11.25.20238725

**Authors:** Harris V. Georgiou

## Abstract

Early vaccination of the general population is a very crucial aspect in the successful mitigation of highly infectious diseases, as it is the case of the SARS-CoV-2 pandemic. The perception of possible side-effects from early batches of vaccines, presumably under-tested, is often a hindering factor for people not in high-risk categories to optin for early vaccination. In this work, early vaccination is formulated under a game-theoretic view with preference ranking and expectation maximization, in order to explore the constraints and conditions under which individuals are keen to opt-in for getting vaccinated. Although simple preference ranking leads to purely non-cooperative / non-altruistic Nash equilibrium, stable cooperative strategies can emerge under simple constraints on the payoffs, specifically the individual cost from possible side-effects versus the collective gain for the community (‘herd’) when endorsing vaccination by default.

**Significance Statement:**

- If the collective gain from community-scale vaccination is deemed even marginally greater than each individual’s cost of possible side-effects, then the best strategy is to ‘cooperate’, i.e., every individual to opt-in for getting vaccinated.
- This condition is independent from the probabilities of getting infected, with or without vaccination, and it is sufficient to lead to a stable cooperative Nash equilibrium.
- In any lethal infectious disease like SARS-CoV-2 and less-than-lethal possible side-effects from the vaccine, for anyone that is susceptible plus one more in his/her own close environment, vaccination is the optimal strategy for all.

## 1. Introduction

One of the most crucial factors in the successful mitigation of the SAR-CoV-2 pandemic is the fast, world-wide adoption of the vaccination plans for the general population. In highly infectious diseases it is imperative that, as soon as a safe vaccine is available, the susceptible portion of the population gets reduced quickly and systematically.

When an adequate proportion of the susceptible population is immunized, the spread of the disease becomes much harder or impossible due to the lack of more carriers. This natural barrier for the infection is often referred to as *herd immunity* and it depends mostly on the infection rate of the disease, more specifically the *basic reproduction number R*_0_, a characteristic property of every infectious disease. Although *R*_0_ varies significantly during an outbreak, simple calculations show that if *R*_0_ is the long-term mean for a specific disease, then herd immunity can be achieved when less than 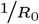 of the population is susceptible, i.e., approximately 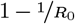 becomes immune either naturally after recovering or via vaccination.

Recent studies on the SARS-CoV-2 have placed its reproduction contagiousness characteristics somewhere between SARS and Ebola viruses. At the early stages of the outbreak its reproduction rate is estimated as 2.5 ≤ *R*_0_ ≤ 3, while at later stages and after the implementation of country-wide lockdown measures it seems to drop to *R*_0_ ≤ 1.7 or even *R*_0_ ≤ 1.4. Thus, a safe threshold of achieving herd immunity for SARS-CoV-2 at its highest rate seems to be somewhere between 0.6-0.7, i.e., 60-70% being naturally (recovered) or artificially (vaccinated) immunized (1).

Perhaps the biggest problem in achieving such a level of long-term protection is the wide adoption of the vaccination plans implemented by each country. On top of the relatively very small fraction of ‘deniers’ of the severity of the pandemic or the disease itself, a valid argument is whether the vaccines that are currently being developed in fast-track processes, with extreme urgency and world-wide demand, will meet all the safety standards and safeguards against possible side-effects, as well as the possibility of lower-than-expected effectiveness. Hence, it is crucial that the overall approach and strategies of the decision-makers on vaccination is examined in a systematic way, so that the proper formulation will highlight the truly important incentives for the societies to adopt such planning. In this brief report, vaccination is modelled under the game-theoretic and probabilistic view in terms of gain-versus-cost for individuals and the ‘herd’ (society), taking into account all the basic factors around disease severity, effectiveness of the vaccine and the possibility of side-effects. The goal is to discover what is the optimal strategy for an individual on the decision to agree or deny vaccination and on what conditions this decision is optimal.

## 2. Vaccination as non-zero sum game

Assuming that all individuals are considered rational players, the decision of participating or not in a voluntary early-vaccination plan can be modelled as a mathematical game (2), more specifically a non-zero sum, since the size of the ‘herd’ at the scale of a whole society (city, region, country) justifies the separation of payoffs. Each person estimates its gains and losses against its own and against the herd, rather than on direct confrontation against other individuals upon the same game value, as it would be the case in the zero-sum case. In other words, every individual must decide the its own gain/loss, as well as the gain/loss for the society, from participating in the early-vaccination plan. The gains are associated with personal immunization and achieving (collectively) herd immunity, while the losses are associated with possible side-effects or ineffectiveness of the vaccine.

There are three main factors to consider for each individual under this formulation:

1. **Vaccination** (Yes/No): Decision to vaccinate or not.
2. **Side-effects** (Yes/No): Resulting side-effects from the vaccine.
3. **Infected** (Yes/No): Infected or not, regardless of vaccination.

From the previous list, an individual is affected by the second and third factor, but can only control the first, i.e., decide whether getting the vaccine or not. It should be noted that no vaccination offers 100% protection against the targeted infection, thus the third factor is valid regardless of this decision, although the probability of infection changes significantly.

Based on these three factors, a mathematical game setup can be described as a sequence of steps and possible outcomes. Table 1 follows the previous enumeration of factors and illustrates the possible combination of outcomes, starting from the decision to vaccinate or not, the possibility of side-effects from the vaccine and finally the infection outcome. It should be noted that getting infected or not is not strictly bounded to either decision of vaccination, thus all possibilities are valid. Evidently, if the no-vaccination route is taken, there is no possibility of side-effects. In total there are six valid outcomes, marked as *c*_0*x*_ for no-vaccination and *c*_1*xx*_ for vaccination.

**Table 1.**
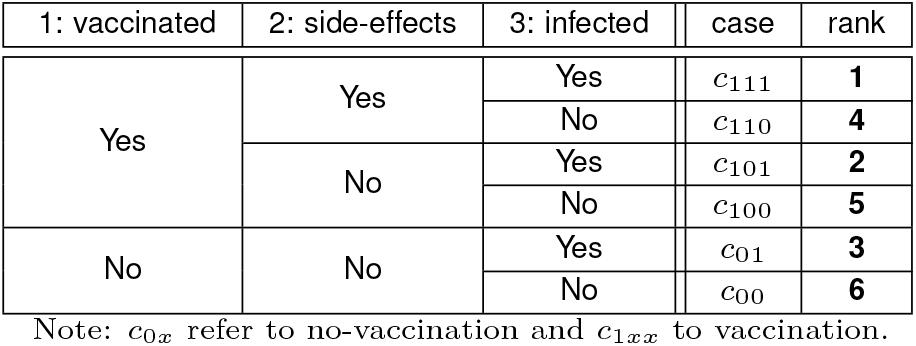
Six possible outcomes based on vaccination, side-effects and infection.

The rightmost column in Table 1 illustrates a typical ranking preference of each outcome, with higher number assigned to more preferable result. For example, 1 is assigned to the case of getting the vaccine, getting side-effects from it and nevertheless getting infected (ineffective). Similarly, 6 is assigned to the case of not getting the vaccine, thus no side-effects, and not getting infected anyway. Intermediate cases may be arranged in different rankings, but the general concept is: (a) not getting infected is always better; (b) not getting the vaccine is preferred if it can be avoided; and (c) if side-effects occur, they are less severe than the infection itself.

From a game-theoretic perspective, Table 1 provides the necessary definition for a N-by-M non-zero sum game setup, where preference ranking is used as individual payoffs. In the simplest case of 2-by-2, the corresponding analytical game is described in Table 2. As expected, the preference ranking of *c*_00_ is the optimal strategy for both players due to the symmetric nature of the game and this outcome is also the Nash equilibrium. In other words, all individuals prefer not to get vaccinated and not getting infected, as expected.

**Table 2.**
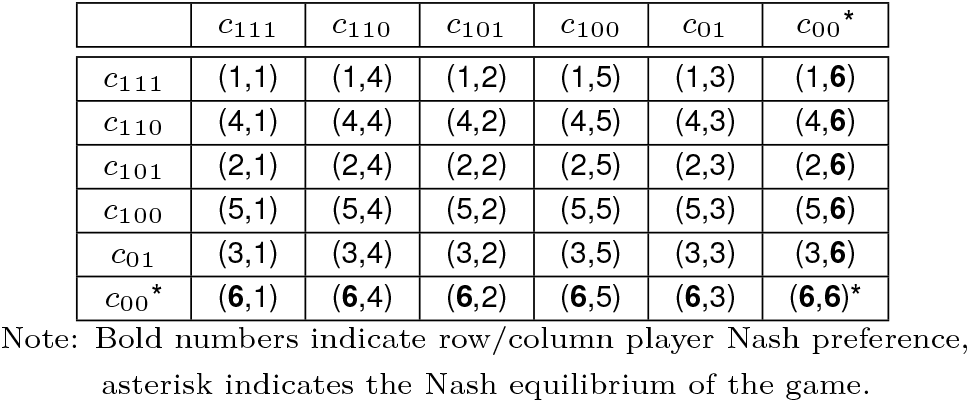
Vaccination 2-by-2 non-zero sum game, using preference ranking as payoffs.

| | $c_{111}$ | $c_{110}$ | $c_{101}$ | $c_{100}$ | $c_{01}$ | $c_{00}^*$ |
| --- | --- | --- | --- | --- | --- | --- |
| $c_{111}$ | (1,1) | (1,4) | (1,2) | (1,5) | (1,3) | (1,6) |
| $c_{110}$ | (4,1) | (4,4) | (4,2) | (4,5) | (4,3) | (4,6) |
| $c_{101}$ | (2,1) | (2,4) | (2,2) | (2,5) | (2,3) | (2,6) |
| $c_{100}$ | (5,1) | (5,4) | (5,2) | (5,5) | (5,3) | (5,6) |
| $c_{01}$ | (3,1) | (3,4) | (3,2) | (3,5) | (3,3) | (3,6) |
| $c_{00}^*$ | (6,1) | (6,4) | (6,2) | (6,5) | (6,3) | (6,6)^* |

Note: Bold numbers indicate row/column player Nash preference, asterisk indicates the Nash equilibrium of the game.

The actual value of this description is to examine how the decision to vaccinate or not groups together the *c*_0*x*_ and *c*_1*xx*_ outcomes. Employing decision *c*_0*x*_, i.e., not getting the vaccine, each player will secure a minimum payoff of *min* {3, 6}= 3, while eploying decision *c*_1*xx*_, i.e., getting the vaccine, each player will secure a minimum payoff of *min* {1, 4, 2, 5}= 1. Using these grouped payoffs, Table 3 illustrates this compact formulation of the game, focusing entirely on the one factor that each player can really control, which is getting the vaccine or not. Still, in this new game the Nash equilibrium is again at the option *c*_0*x*_, i.e., not getting the vaccine. However, the game matrix is now very similar to two very important game setups in Game Theory, namely the Chicken game (CK) and the Prisoners’ Dilemma (PD) (2). In both cases, the ‘winning’ condition in the asymmetric pairs needs to exhibit payoff larger than any other case, while the difference between CK and PD becomes evident when the corresponding ‘loss’ is better (CK) or worse (PD) than when both players loose.

**Table 3.** Vaccination 2-by-2 non-zero sum game, using grouped preference ranking as payoffs.

| | $c_{1xx}$ | $c_{0x}$ |
| --- | --- | --- |
| $c_{1xx}$ | (1,1) | (1,3) |
| $c_{0x}$ | (3,1) | (3,3)^* |

Note: Bold numbers indicate row/column player Nash preference, asterisk indicates the Nash equilibrium of the game.

The vaccination game of Table 3 is transformed into CK if getting the vaccine while the others do not is a loss, but not as much as if no one gets the vaccine, assuming that any side-effects from it is much less severe than everyone getting infected as a result of no-vaccination. Similarly, the vaccination game Table 3 is transformed into PD if getting the vaccine while the others do not is the most severe loss, even when compared to everyone getting infected as a result of no-vaccination. More details in the four most important non-zero sum games can be found in (3).

In order to examine the conditions under which the vaccination game becomes CK or PD or any other setup with non-trivial solution, the preference ranking must be re-defined in a more analytical formulation via probabilistic expectancies with regard to gain/loss from each outcome. This alternative approach is examined next in section 3.

## 3. Vaccination as expectation maximization

The preference ranking in Table 1 is a valid approach in Game Theory in order to get a general perspective of a game setup and outcomes, assuming that all players have fixed strategy preferences (mixed-strategy probabilities) and associated pay-offs. In reality, what a player typically wishes to do is to estimate an optimal *mixture* of strategies for equal expected payoffs regardless of the opponents’ choices, rather than select a single strategy. Thus, a player may actually allocate its choices probabilistically between e.g. preference 5 and 6, so that both strategies are valid for selection in an optimal way.

Based on the three main factors of the vaccination game as presented in section 2, there are a few parameters that can clearly define the setup in terms of probabilistic expectancy and associated payoffs.

Regarding the probabilities of outcomes, these are:

- 0 ≤ *p*_00_ 1 ≤ : Probability of no infection, without vaccination.
- 0 ≤ *p*_10_ ≤ 1: Probability of no infection, with vaccination.
- 0 ≤ *p*_*s*_ ≤ 1 : Probability of side-effects from vaccination.

Regarding the associated payoffs, these are:

- *α*_0_ ≥ 0 : Individual gain when not infected.
- *γ*_1_ ≥ 0 : Group gain when vaccinated.
- *β*_*s*_ ≥ 0 : Cost(-) of vaccination side-effects for the individual.

Given these definitions and using the complementary probabilities (1 − *p*) for the opposite outcomes, Tables 4 and 5 provide an analytical definition of probabilities and payoffs for all cases *c*_*xxx*_, i.e., with and without vaccination.

**Table 4.** Probabilities for each outcome from choices *c*_*xxx*_.

| | not vaccinated ($c_{0x}$) | vaccinated ($c_{1x}$) |
| --- | --- | --- |
| $c_{x00}$ | $p_{00}$ | $(1 - p_s) \cdot p_{10}$ |
| $c_{x01}$ | $(1 - p_{00})$ | $(1 - p_s) \cdot (1 - p_{10})$ |
| $c_{x10}$ | 0 | $p_s \cdot p_{10}$ |
| $c_{x11}$ | 0 | $p_s \cdot (1 - p_{10})$ |

Note: $c_{xxx}$ are as defined in Table 1; $0 \leq \{p_{00}, p_{10}, p_s\} \leq 1$ .

**Table 5.** Payoffs for each outcome from choices *c*_*xxx*_.

| | not vaccinated ($c_{0x}$) | vaccinated ($c_{1x}$) |
| --- | --- | --- |
| $c_{x00}$ | $\alpha_0$ | $\alpha_0 + \gamma_1$ |
| $c_{x01}$ | 0 | $\gamma_1$ |
| $c_{x10}$ | $\alpha_0 - \beta_s$ | $\alpha_0 - \beta_s + \gamma_1$ |
| $c_{x11}$ | $-\beta_s$ | $-\beta_s + \gamma_1$ |

Note: $c_{xxx}$ are as defined in Table 1; $\{\alpha_0, \beta_s, \gamma_1\} \geq 0$ .

Using the definitions in Tables 4 and 5, the expected payoffs for an individual player can be analytically defined for both choices, i.e., getting vaccinated or not. Specifically, for each choice there are four outcomes for which the specific payoff is combined with the associated probability. For not getting vaccinated these are *ξ*_*i*_, *i* ={1, …, 4}, and for getting vaccinated these are *ϕ*_*i*_, *i* ={1, …, 4}, as defined in the Eq.1-4 and Eq.5-8, respectively.

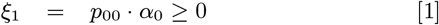

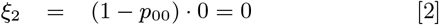

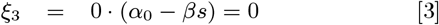

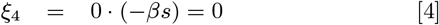

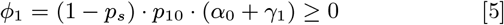

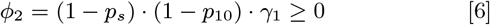

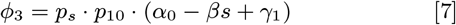

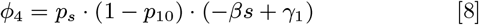

In the sense of *minimax* optimality from Game Theory, the player wishes to select the maximum of the minimum expected payoffs from each strategy, i.e., select the choice that leads to the largest guarantee regarding the payoff. This means that the minimum is selected for each of the *ξ*_*i*_ and *ϕ*_*i*_ sets and then the maximum of the two resulting payoffs dictate the largest-expected-payoff strategy regarding whether to vaccinate of not. Hence, for the first step (minimums) these are:

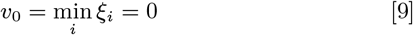

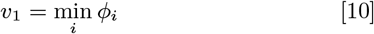

and for the second step (maximum) its is:

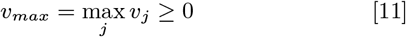

It is worth noting that for *v*_0_ the minimum can be easily defined as zero, since it is evident from Eq.2-4. This translates to the ‘threshold’ of minimum expected payoff from not getting vaccinated, which is what the choice of getting vaccinated should surpass in order to be dictated as optimal, i.e.:

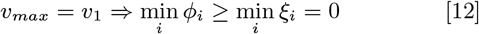

From the definitions of payoffs in Table 5 it is clear that *ϕ*_1_ ≥ 0 and *ϕ*_2_ ≥ 0. Hence, the only two options for less-than-zero payoffs that may lead to smaller minimum are the ones that involve the cost −*β*_*s*_ of possible vaccination side-effects, i.e., *ϕ*_3_ and *ϕ*_4_. Subsequently, for getting an expected payoff from {*ϕ*_3_, *ϕ*_4_} (with side-effects) no worse than from {*ϕ*_1_, *ϕ*_2_} (no side-effects) the corresponding constraints are:

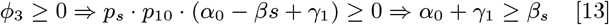

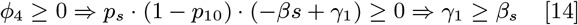

and according to the definitions in Table 5 the constraints of Eq.13-14 can be combined as:

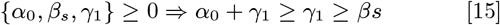

## 4. Discussion

The rightmost inequality in Eq.15 is the crucial factor for defining the optimal strategy between *ϕ*_*i*_ and *ξ*_*i*_, i.e., getting vaccinated or not, respectively:

*If the collective gain γ*_1_ *from vaccination is deemed even marginally greater than the individual cost β*_*s*_ *of possible side-effects, then the best choice of every individual is to get vaccinated*.

Intuitively, this outcome is what is expected in every vaccination plan for any infectious disease. That is, if the possible side-effects are deemed less severe than the community-scale damage from the disease, then the ‘herd’ is better off endorsing the vaccination for the entire community or at least at the ‘herd immunity’ threshold of 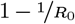, as described earlier in section 1. Although there is always the risk of ‘free-riders’ as in every collaborative gaming, strategies that include both ‘kindness’ and reciprocity like ‘tit-for-tat’ have been proven as sufficient for the emergence of Evolutionary Stable Strategies (ESS) that are cooperative (4). Practically, in any lethal infectious disease like SARS-CoV-2 and less-than-lethal possible side-effects from the vaccine, for anyone that is susceptible plus one more in his/her own close environment, vaccination is the optimal strategy.

Furthermore, it is very interesting to see that this constraint is independent from the probabilities of getting infected with (1 − *p*_10_) or without (1 − *p*_00_) vaccination. This is because the probability *p*_*s*_ and cost *β*_*s*_ of side-effects are taken into account for both cases of getting infected or not after vaccination. In other words, the only thing that defines the minimum from *ϕ*_*i*_ in Eq.5-8 is comparing the expected payoffs with and without side-effects from vaccination. This is perhaps the most important result from the expectation maximization analysis presented here.

## 5. Conclusion

In this work, early vaccination is formulated under a game-theoretic view, first with preference ranking and subsequently with expectation maximization, in order to explore the constraints and conditions under which individuals are keen to opt-in for getting vaccinated. Preference ranking leads to purely non-cooperative / non-altruistic Nash equilibrium, not much differently than in Chicken or Prisoners’ Dilemma games. Nevertheless, as in these games too, cooperative strategies can emerge as stable Nash equilibria under simple constraints on the payoffs, specifically the individual cost from possible side-effects versus the collective gain for the community (‘herd’) when endorsing vaccination by default. It is very important for policy-makers to understand and communicate these aspects promptly and effectively, in order to implement successful early vaccination plans as in the case of SARS-CoV-2.

## Data Availability

No datasets were used in this study.

## ACKNOWLEDGMENTS

The author wishes to thank every team and researcher that is currently working towards scientific works of high quality, readily accessible to the community, including technical papers, research papers and datasets. This is the only viable way to address such fast-pacing events of major world-wide impact collectively and effectively, as quickly and reliably as possible, while the crisis is still evolving.

